# Early identification of Family Medicine residents at risk of failure using Natural Language Processing and Explainable Artificial Intelligence

**DOI:** 10.1101/2024.12.07.24318566

**Authors:** Abhisht Makarand Joshi, Pouria Mortezaagha, Diana Inkpen, Edward Seale, Douglas Archibald, Kendall Noel, Arya Rahgozar

## Abstract

**Background:** During residency, each resident is observed and receives feedback based on their performance. Residency training is demanding, with some residents struggling with their academic performance. A competency-based residency training program’s success depends on its ability to identify residents with difficulty during their first year of post-graduate education and to provide them with timely intervention and support.

**Objective:** In large training programs such as Family Medicine, identifying residents at risk of failing their certification exams is difficult. We developed an AI system using state-of-the-art technologies in Machine Learning (ML), Deep Learning (DL), Natural Language Processing (NLP) and Explainable AI (XAI) to detect at-risk residents automatically.

**Materials and Methods:** The research was conducted in the 2023-24 academic year. We implemented ML, DL and NLP models for prediction and performance analysis. The target variable chosen for the prediction was the determination of whether the resident would fail or pass their certification exam. XAI was used to enhance the understanding of the model’s inner workings.

**Results:** In total, there were 1382 data points of residents. The final model, Support Vector Machine (SVM), achieved an accuracy of 89.05% and an F1 score of 74.54 for the multiclass classification when multimodal (text and tabular) data was used. This model outperformed the models that only used qualitative or quantitative data exclusively.

**Conclusion:** Combining qualitative and quantitative data represents a novel approach and provided better classification results. This research demonstrates the feasibility of an automated AI system for the early identification of residents at risk of academic struggle.

**Prior Abstract Presentation:** Abstract presented at AMEE (An International Association for Medical Education) Conference Basel, Switzerland, August 24-28, 2024.

## Introduction

Residency programs play a crucial role in molding future specialist and generalist physicians by providing them extensive clinical experience. When residents encounter problems late in their residency, it is often more difficult to rectify them. Ideally, these problems should be identified early in residency, which would allow them to be resolved promptly, avoiding ramifications, such as failure on certification examinations. By Early we mean, as soon as the resident evaluation data becomes available. Early identification of at-risk residents is a vital responsibility of a residency’s program of evaluation. Artificial intelligence (AI) is effective for early detection because it can analyze large volumes of data quickly, identifying subtle patterns and risk factors that may be missed by human evaluators, enabling timely interventions. Thus, AI is suggested to be used for *early* identification of at-risk residents. By *early* we mean, as soon as the resident evaluation data becomes available.

According to a survey of internal medicine program directors, struggling residents display characteristics of inadequate medical knowledge, poor clinical judgment, and inefficient use of their time^1^. Additionally, struggling residents may feel overburdened, unsure of the objectives of training, unclear about their performance evaluation and thus incapable of prioritizing areas of improvement^2^. The demanding nature of residency programs, the long work hours, and the stress of transitioning to residency, all play a role in these challenges^3^.

The use of Machine Learning (ML) and Natural Language Processing (NLP) in medical education to improve students’ and healthcare professionals’ teaching and learning processes has gained popularity in recent years. Several promising strategies have emerged because of studies exploring the potential applications of NLP^4–10^ and ML^11–19^ in medical education. This emphasis on NLP has practical applications. Automated essay scoring (AES) for medical knowledge examinations and constructed-response assignments, automates grading by linking language to human scores using word processing and NLP^4,5^. NLP was applied to examine faculty and medical residents’ feelings about entrustable professional activity (EPA) evaluations revealing that general surgery participants expressed fewer positive emotions compared to those in emergency medicine^6^. Explainable AI (XAI) has shown to improve decision making and educational results, by improving a prediction model’s transparency and comprehension^20–24^. The explainability of the otherwise complex reasons behind ML predictions play an important role in establishing end users’ trust in AI systems’ credibility, by mapping the important patterns, phrases and terms associated with the highest information value for the predictions^25^. Few studies have explored the role of Large Language models (LLMs) like ChatGPT in medical education. ChatGPT’s usage in anatomical education was examined, stressing its potential for tailored learning but criticizing its errors, academic exploitation, and limited visual processing skills, therefore limiting its dependability as a standalone tool^26^. Another study investigated the use of GPT^a^ to create clinical assessment items, finding comparable quality to standard methods with reduced cost^27^.

The identification of at-risk residents and the subsequent educational interventions can be greatly improved with the use of AI. We propose a multimodal approach of combining Qualitative (Text) and Quantitative (Numerical) data utilizing Post Graduate Year 1 (PGY 1) Family Medicine rotation data, along with XAI for model explanation. This integration of data in the form of text and narration, allows AI systems to consider real-life experiences^28^. Integration of humanistic opinions can enhance AI’s data-centric methods. Our study attempts to close the research gap, by investigating the efficiency of combining qualitative and quantitative data, via three independent sets of experiments. By examining the synergy between the two types of data, we hope to show that their combination produces more valid results. The study makes use of advanced models such as XLNET^29^ for the prediction of the residents at risk, along with the implementation of XAI to help improve the end user’s understanding. *Figure 1* displays the intuitive approach for the model.

**Figure 1.**
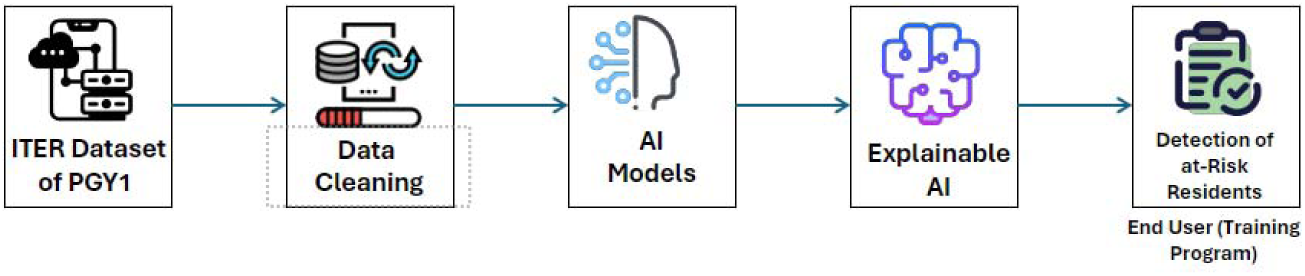
System Architecture for the experiment

The objectives of our study are twofold. First, we will explore the feasibility of automating the identification and prediction of at-risk residents in educational trouble using ML and NLP. Second, we will use XAI and LLMs, to identify the important latent characteristics, discriminating patterns and generate insights.

## Materials and Methods

### Research Setting and Participants

The dataset utilized for the study is the Family Medicine In-Training Evaluation Report (ITER) of first-year residents (PGY-1) and their certification exam data. The ITER data is sourced from the Department of Family Medicine at the University of Ottawa. The exams data is based on the CFPC^b^ certification exam, which has two components Simulated office oral (SOO) and Short-answer management problems (SAMP) and they make up the Canadian Family Medicine Certification exam. The exams are offered in the Spring and the Fall of each year. The exam data used covers the following periods: Spring: 2018-2022, Fall: 2018 - 2022. There was no data for Spring 2020, as the exam was cancelled due to the COVID-19 pandemic.

This study was conducted in the year 2023-24. We focused on analyzing the performance outcomes represented by the target variable “Pass/Fail,” which is a multiclass classification with four different classes, Fail in SOO, Fail in SAMP, Fail in both and Pass (See *Figure 2*). The dataset is imbalanced^c^, meaning some classes have a much greater count than others. Exam results are recorded as z-scores for the SOO and the SAMP where a z-score of less than −2.0 marks a failure. Of the 1382 SOO scores, 61 fell below this level. Of 1382 SAMP scores, 21 fell below −2.0. Only 8 out of 1382 failed both parts. Most candidates passed: 1292 out of 1382 either passed both components or one if only one was available. This distribution shows a significant skew towards the “PASS” category, highlighting the imbalance in the dataset.

**Figure 2.**
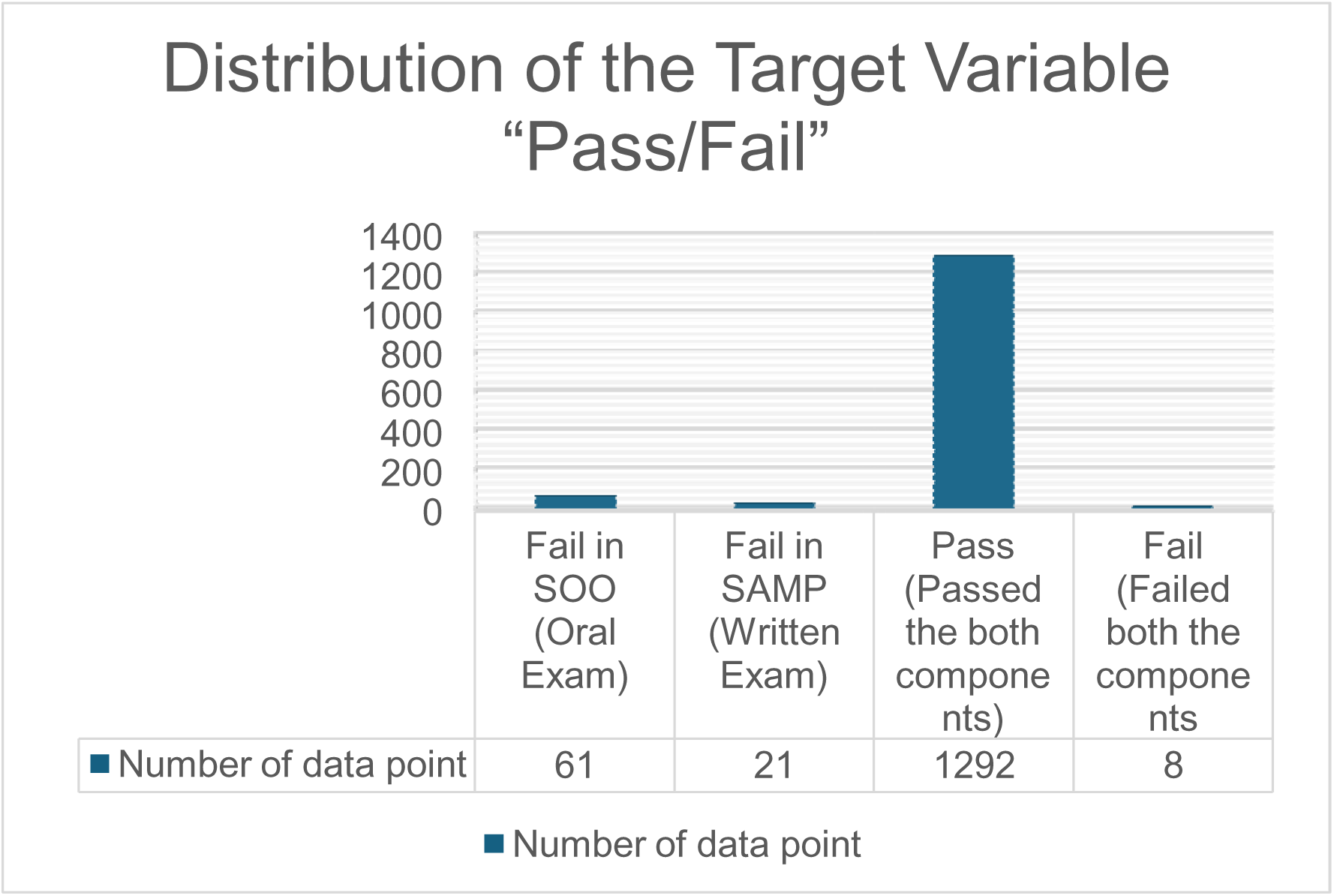
Distribution of Target Variable “Pass/Fail”

### Data Availability Statement

Due to the sensitive nature of this data, which includes confidential information about medical residents, it cannot be publicly shared. Access to the data is restricted to ensure participant privacy and confidentiality.

### Interventions

This section presents the techniques employed to develop an AI-based medical residency intervention system. *Figure 3* illustrates the comprehensive approach implemented in our methodology.

**Figure 3.**
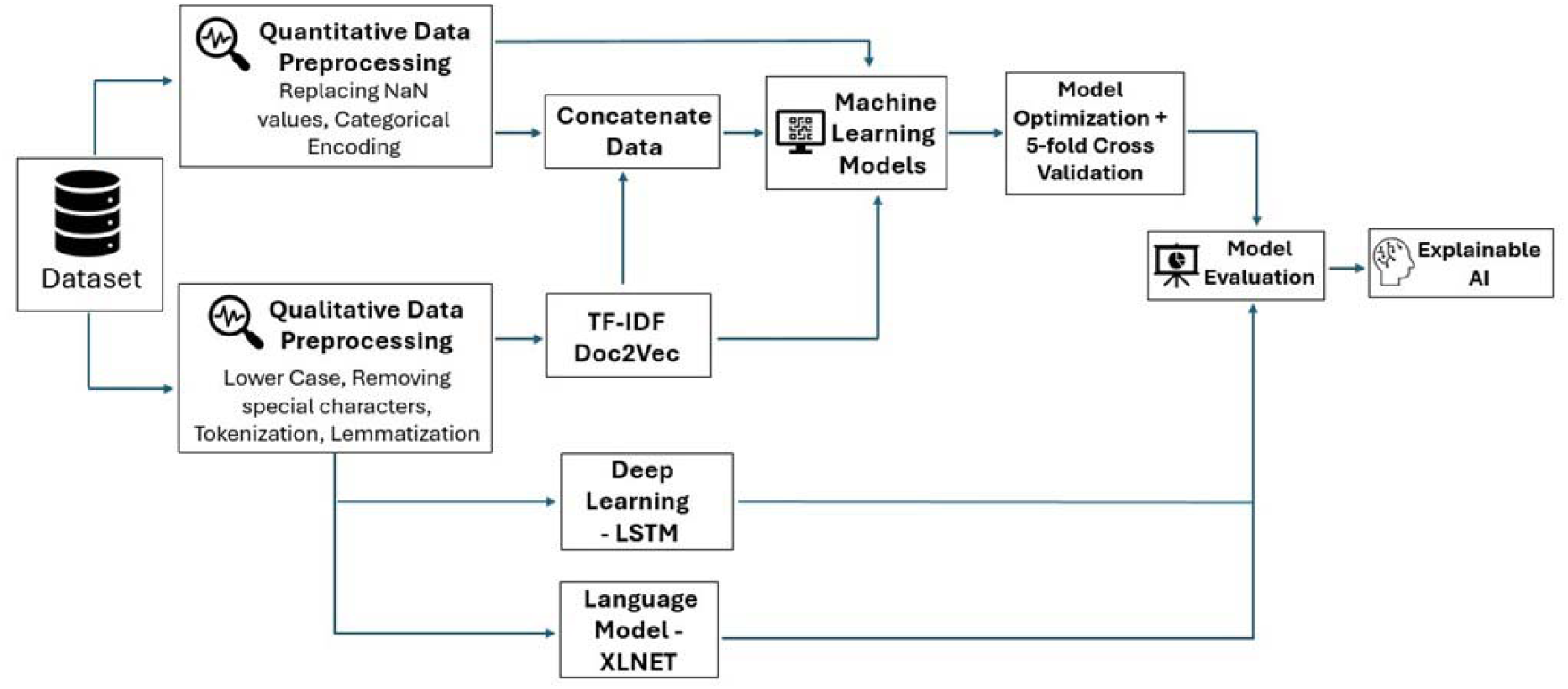
Research Methodology for Medical Residency Intervention

The dataset comprises 47 input columns, of which nine features were qualitative requiring narrative comments, and the remaining thirty-eight features were quantitative evaluations of the residents. This data was constructed from the assessment conducted by faculty members based on the resident’s performance during their family medicine rotations in residency during their first year. We aim to utilize a multimodal approach, which involves combining quantitative (tabular) and qualitative (text) data, which includes feedback, and comments offered by the faculty to each resident. Furthermore, the examined dataset also includes the residents’ final Pass or Fail status on their first certification exam attempt.

### AI Modelling

Data were cleaned in preparation for the experiment. The cleaning process involved filling empty entries and performing categorical encoding to the quantitative part of the data. Subsequently, a multi-step cleaning process was implemented for the qualitative data, encompassing converting the text to lowercase, removing special characters, tokenizing, and lemmatizing the text. We then vectorized the data using TF-IDF^30^ to convert the text into numerical representations and produced dense vector representations encapsulating the semantic meaning of the text using Doc2Vec^31^ embeddings for additional model processing. A Language Model XLNET and a Deep Learning model Long Short-Term Memory (LSTM)^32^ which is a type of Recurrent Neural Network designed to remember long-term dependencies were applied on qualitative data. You can find the link to the GitHub repository^d^ for the project in the footnote, which outlines the basic code and implementation details of this project.

### Outcomes Measured

Our study involved a series of three AI experiments utilizing “Pass/Fail” as the target variable for all the three experiments. For comparison purposes, we experimented with three kinds of data within the dataset: qualitative, quantitative and multimodal.

#### I. Series of Experiments I: Qualitative Data

In these experiments, we employed four approaches focusing solely on qualitative data via a) ML models on text vectorization (TF-IDF), b) ML Models on text embeddings (Doc2Vec), c) fine tuning the pre-trained XLNET on the data and d) LSTM, a Deep Learning model.

#### II. Series of Experiments II: Quantitative Data

We utilized only quantitative data. SMOTE^33^ was applied to balance the data. We used SMOTE to synthetically increase the short segments within the dataset without introducing any bias. The experimental methodology followed these steps: feature scaling and normalization, Principal Component Analysis (PCA), feature selection (Mutual Information gain), hyperparameter tuning (Grid search) and model evaluation (Cross-validation) and prediction.

#### III. Series of Experiments III: Multimodal (Combination of Qualitative and Quantitative Data)

This final series of experiments aimed to enhance information richness and evaluate whether multimodality benefits the AI model’s performance. Before concatenating the data, we applied TF-IDF and Doc2Vec to text data, followed by SMOTE, as done previously. Finally, we plugged the transformed data into the algorithm.

### Analysis of Outcomes

#### Evaluation Metrics and Cross Validation

The F1 (macro) score and accuracy were selected as evaluation metrics to measure model performance (*See Supplementary Material 1*). A 5-fold cross-validation was performed on the stratified data of which four parts were used for training the AI model and one part for testing. Through the cross-validation we reported the average of the indices with a variance of up to 3%^34^.

### Explainable AI (XAI)

Explaining an AI model means rendering its output understandable to a human being^35^. XAI plays a crucial role in understanding what is going on behind a complex ML model. To gain insights into our AI model, SHAP^36^ and BERTopic^37^ were employed for model explanation and to understand why certain predictions were made. SHAP helped us identify important input variables, akin to the industrial term “fine classing,” which is comparable to the PCA (Principal Component Analysis) used in our process. On the other hand, BERTopic is a topic modelling technique that leverages language model and assists in “explaining” output predictions by identifying and grouping topics within the textual data. Our utilization of BERTopic as a tool for “Global Explainability” is reinforced using a Language Model (XLNET). The key features were then shared with an expert.

### Shapley Additive Explanations (SHAP)

SHAP was used for model interpretation to identify crucial features and provide their order of importance. Together this information would allow the faculty and supervisors to understand which aspects of the residency to focus on when a resident is at risk.

### BERTopic

BERTopic, is a cutting-edge topic modelling technique that leverages BERT^38^ embeddings and class-based TF-IDF to create dense clusters, enabling the generation of easily interpretable topics while retaining important words in the topic descriptions. By integrating BERT embeddings, UMAP for dimensionality reduction, and HDBSCAN for clustering. BERTopic facilitates the identification of distinct topics within a corpus of text data. This will help the end user capture necessary information and contextual understanding using a state-of-the-art model for accurate and rich topic presentation. This approach is particularly valuable in the context of XAI, as it emphasizes transparency and interpretability in the modelling process.

### Feasibility and Acceptability

These models were feasible in time and resources, making them an effective screening tool. Sharing key features with experts ensured feasibility, relevance, and acceptability.

### REB statement

The project’s scope was reviewed by the Office of Research Ethics and Integrity at the University of Ottawa and that this project falls within Article 2.5 of the TCPS^e^ 2 was determined deemed exempt from further review.

## Results

We reported the score averages of performance of the ML models for three sets of experiments.

### I. Series of Experiments I: Qualitative Data

For Experiment 1, we used the following models: *Multinomial Logistic Regression (MLR), Bernoulli Naïve Bayes(BNB), Gaussian Naïve Bayes(GNB), SVM, LSTM* and *XLNET*. *XLNET* outperformed the other models with an accuracy of 72.45% and an F1 score of 55.48. *LSTM* performed second best with an accuracy of 70.33% and an F1 score of 53.73. Table 1 shows the model performance for each model in Experiment 1.

**Table 1.**
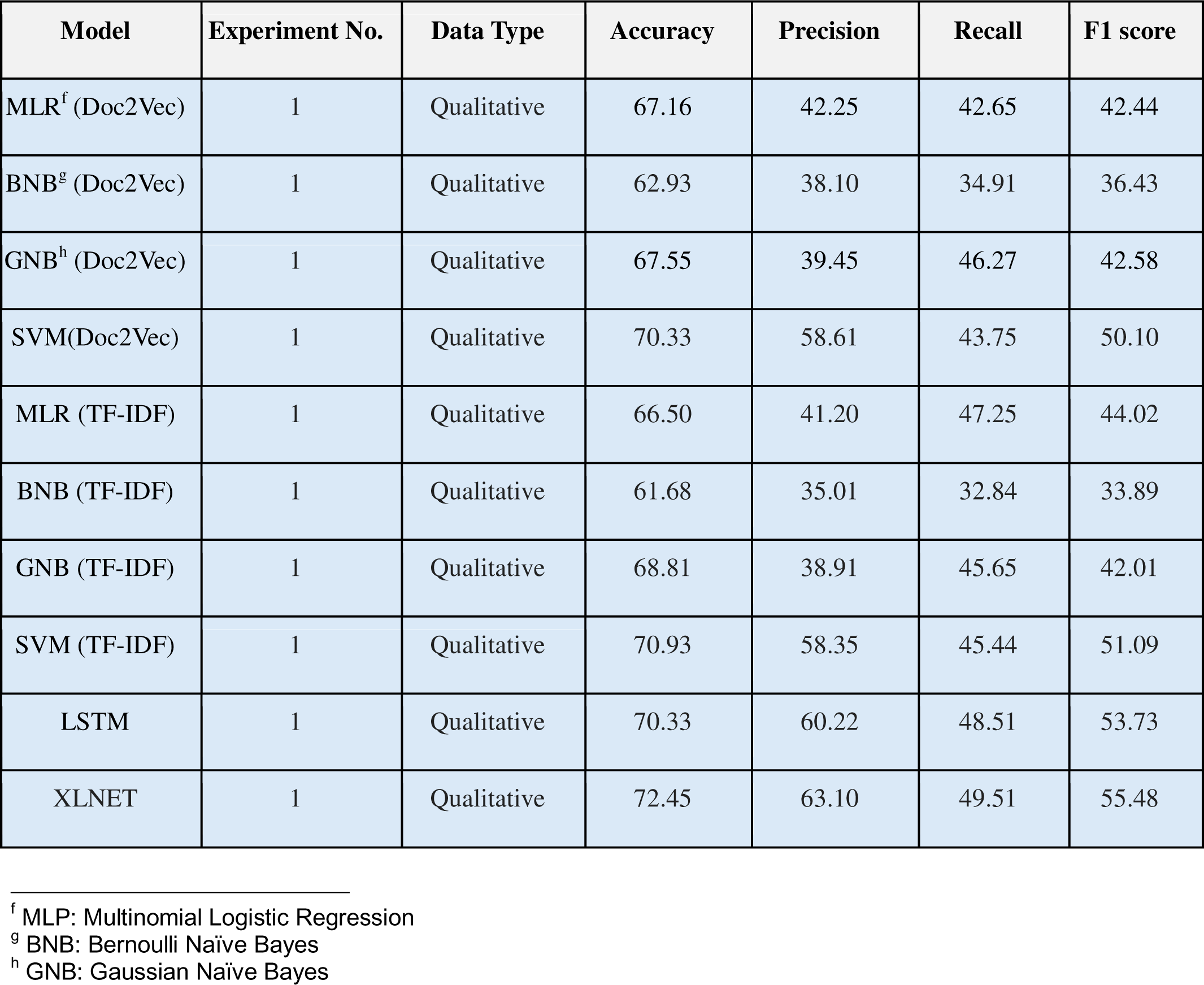
Model Performances of Experiment 1.

### II. Series of Experiments II: Quantitative Data

In Experiment 2, the following ML models were utilized, *SVM, Random Forest, Multilayer Perceptron and CatBoost*. The *SVM* model, followed by the *CatBoost* model, achieved the highest performance with an accuracy of 81.71% and 80.93%, and an F1 score of 63.43 and 63.01, respectively. Table 2 shows the model performance for each model in Experiment 2.

**Table 2.** Model Performances of Experiment 2.

|  | Experiment No. | Data Type | Accuracy | Precision | Recall | F1 score |
| --- | --- | --- | --- | --- | --- | --- |

<sup>f</sup> MLP: Multinomial Logistic Regression
<sup>g</sup> BNB: Bernoulli Naïve Bayes
<sup>h</sup> GNB: Gaussian Naïve Bayes

| Model |  |  |  |  |  |  |
| --- | --- | --- | --- | --- | --- | --- |
| SVM | 2 | Quantitative | 81.71 | 64.98 | 61.97 | 63.43 |
| RF<sup>i</sup> | 2 | Quantitative | 76.43 | 62.60 | 59.10 | 60.79 |
| MLP<sup>j</sup> | 2 | Quantitative | 77.92 | 64.04 | 59.79 | 61.84 |
| CatBoost | 2 | Quantitative | 80.93 | 66.10 | 60.94 | 63.01 |

**Table 3.** Model Performances of Experiment 3.

| Model | Experiment No. | Data Type | Accuracy | Precision | Recall | F1 score |
| --- | --- | --- | --- | --- | --- | --- |

<sup>i</sup> RF: Random Forest
<sup>j</sup> MLP: Multi-layer Perceptron

|  |  |  |  |  |  |  |
| --- | --- | --- | --- | --- | --- | --- |
| MLR (Doc2Vec) | 3 | Combined | 75.42 | 69.48 | 67.85 | 68.65 |
| BNB(Doc2Vec) | 3 | Combined | 70.79 | 52.50 | 49.85 | 51.14 |
| GNB (Doc2Vec) | 3 | Combined | 77.60 | 71.80 | 69.85 | 70.81 |
| SVM(Doc2Vec) | 3 | Combined | 82.10 | 74.80 | 70.15 | 72.40 |
| MLR (TF-IDF) | 3 | Combined | 80.26 | 73.45 | 70.10 | 71.73 |
| BNB (TF-IDF) | 3 | Combined | 70.24 | 54.22 | 48.45 | 51.17 |
| GNB (TF-IDF) | 3 | Combined | 82.15 | 72.32 | 69.69 | 70.98 |
| SVM (TF-IDF) | 3 | Combined | 89.05 | 76.11 | 73.04 | 74.54 |

### III. Series of Experiments III: Multimodal Data

For Experiment 3, the following ML models were utilized, *MLR, BNB, GNB* and *SVM*. The *SVM (TF-IDF)* model outperformed all other models with an accuracy of 89.05% and F1 score of 74.54, followed by *SVM (Doc2Vec)* with an accuracy of 82.10 and an F1 score of 72.40.

The results clearly indicate that the *SVM (TF-IDF)* model from Experiment III was the champion. It used TF-IDF on text data and a multimodal approach, outperforming other models in Experiments I, II, and III. Interestingly, despite Doc2Vec being context-dependent, the TF-IDF model outperformed it, likely due to TF-IDF’s better performance with smaller datasets^39^.

### Explainable AI

#### SHAP

The summary plot for the most important features is shown in *Figure 4* and is explained below: The summary plot shows the feature in order of importance, where the bar length for each class represents its impact on that class. Target variables were labelled as follows, Class 0 for students who fail SAMP, Class 1 for those who fail SOO, Class 2 for those who fail both, and Class 3 for students who pass both SAMP and SOO. Below are explained the six most important features according to SHAP and comments of the expert (Supervisor of the residents).

1. **Feature 1 (pc0):** The most crucial feature was, *“Were the rotation objectives discussed with the resident?”* This feature has two possible answers: Yes or No. It was observed that the residents who had discussed their objectives with their supervisor had a failure rate of 7.2%, but those who did not, had a failure rate of just 4.1%. It is hypothesized that during residency, there is a higher chance that the rotation’s objectives will be discussed if the resident is having difficulty. This contrasts with a resident who is known or perceived to be doing well and who has not undertaken these measures with the faculty.
2. **Feature 2 (pc1)**, “*Was a learning plan used?”* The question has 2 possible answers. No - 1 and Yes - 2. As per above 6.6% of residents with NO learning plan failed the exam, while 9.2% of those who did have one failed. In a world where learning plans are for the most part only used and/or talked about when a learner is having difficulties, it perhaps makes sense that a greater percentage of them fail the exam. However, in the future, we hope to encourage all learners to have a learning plan (a philosophical debate), so that it may become a moot point.
3. **Feature 3 (pc3)**: *Begins to engage in scholarly activity (e.g. creation, dissemination, application & translation of knowledge by completing a scholarly project / practice audit / quality improvement process)*. (Numerical Answer). This is a question with 4 possible answers:

0 - Not observable or not applicable = 5.7% (8/141) failed.
1 - Off trajectory for this PGY1 benchmark (action required) = 0% (0/4) Failed.
2 - On trajectory for this PGY1 benchmark (minimal or no action required) = 8.2% (52/629) Failed.
3 - Attained this PGY1 Benchmark (no action required) = 4.9% (30/608) Failed
4. **Feature 4 (pc30):** Uses patient centered interviewing principles to obtain / demonstrate understanding of the context of the “whole person” & find common ground in management. (e.g. Feelings, Ideas, effect on Function, Expectations to understand both the patient’s illness & illness experience; develops contextual appreciation of the patient’s family, education, employment, finances, religion & supports.) (Numerical Answer). This is a question with 4 possible answers:

0 - Not observable or not applicable = 0% (0/3) failed.
1 - Off trajectory for this PGY1 benchmark (action required) = 33.3% (2/6) Failed.
2 - On trajectory for this PGY1 benchmark (minimal or no action required) = 8.0% (47/588) Failed.
3 - Attained this PGY1 Benchmark (no action required) = 5.2% (41/785) Failed This is a crucial skill for the oral exams, so to see a higher rate of percentage with the lower scores makes sense.
5. **Feature 5 (pc10):** Demonstrates comfort with diagnostic uncertainty inherent to family medicine, such that clinical assessment is appropriately selective. (Numerical Answer). This is a question with 4 possible answers:

0 - Not observable or not applicable = 0% (0/5) Failed
1 - Off trajectory for this PGY1 benchmark (action required) = 26.3% (5/19) Failed
2 - On trajectory for this PGY1 benchmark (minimal or no action required) = 6.9% (54/779) Failed
3 - Attained this PGY1 Benchmark (no action required) = 5.3% (31/579) Failed This is another very important skill. It is nice to see that a higher score appears to be correlating to a lower fail rate and that it is among the predictive features.
6. **Feature 6 (pc2):** Accesses & evaluates medical literature, interpreting articles using principles of evidence-based medicine. (Numerical Answer). This is a question with 4 possible answers:

0 - Not observable or not applicable = 0% (0/189) Failed
1 - Off trajectory for this PGY1 benchmark (action required) = 0% (0/5) Failed
2 - On trajectory for this PGY1 benchmark (minimal or no action required) = 8.1% (49/607) Failed
3 - Attained this PGY1 Benchmark (no action required) = 5.5% (32/581) Failed This is another very important skill. It is nice to see that a higher score appears to be correlating to a lower fail rate and that it is among the predictive features.

**Figure 4.**
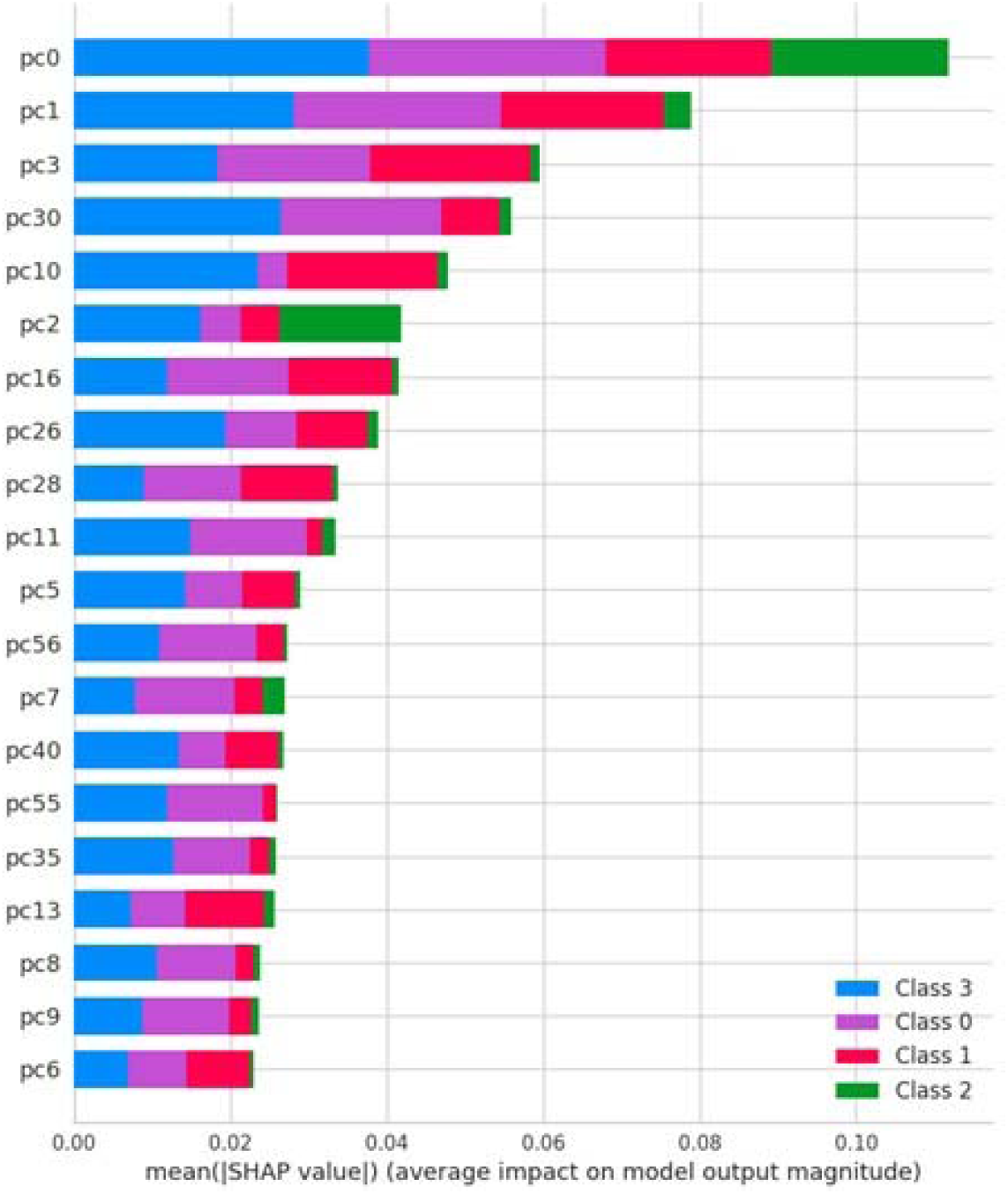
SHAP Summary plot

A final comment was also offered by the expert regarding the overall interpretation of the model, which was as follows: “*Features 4 and 5 were reassuring, while features 3 and 6 were more surprising.*”

### BERTopic

BERTopic has the potential to produce several topics, for the sake of clarity we have selected the top seven topics. *Figure 5* presents the top terms for each topic using a bar chart that shows the importance of each word for the topic. The horizontal axis is the c-TF-IDF scores of the top five most representative words for each topic. For instance, Topic 1 *“Trajectory”* is related to the progress of the medical resident during their residency and their supervisor comments on whether the resident is off-trajectory or on-trajectory. It was observed that residents who were on-trajectory had a lower failure rate compared to the ones who were off-trajectory. Topic 2 *“Communication”* and Topic 3 *“Notes”* for instance, those residents who have positive feedback on their communication and timely note taking and documentation had passed the certification while those who had comments on improving the note taking and communication failed in the certification exams with a few exceptions in both the cases. However, timely and correct note taking, and good communication can be an indicator of the resident’s performance.

**Figure 5.**
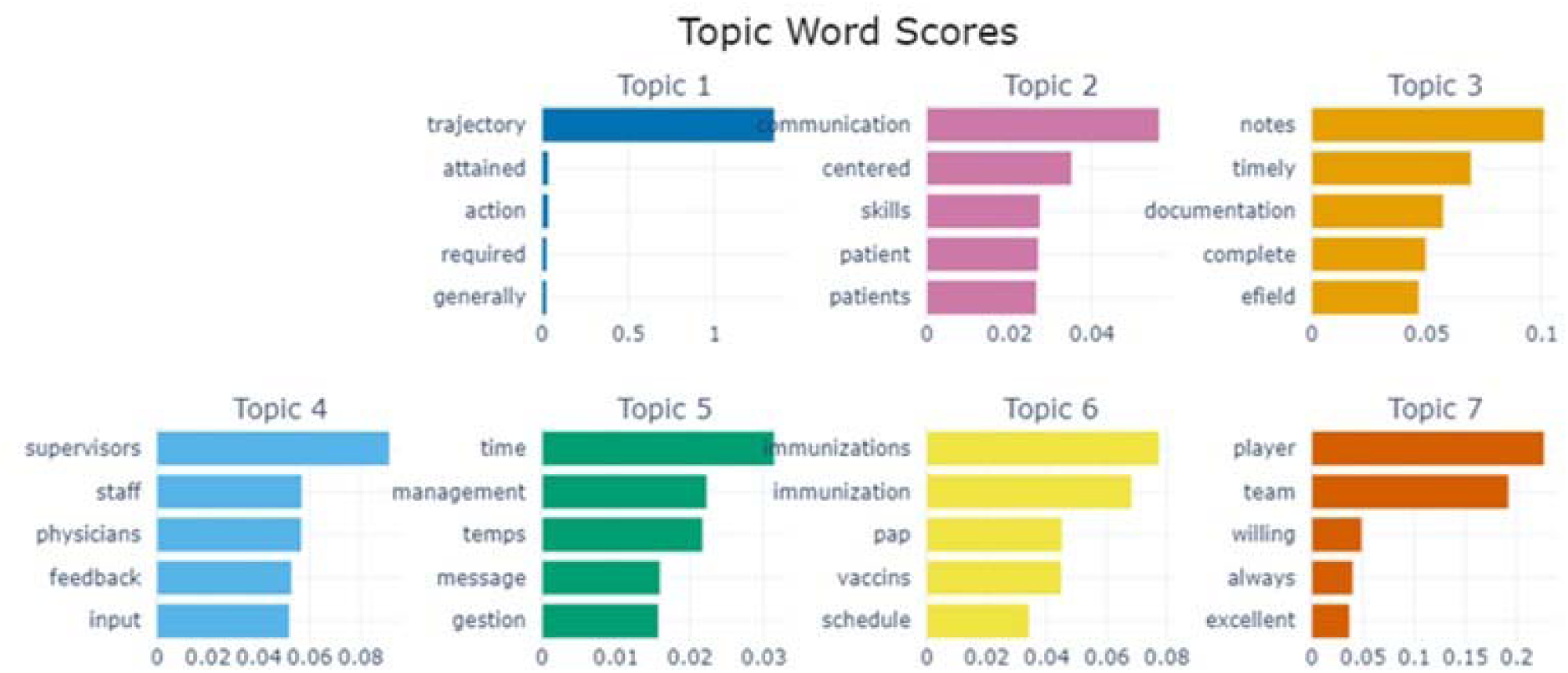
Topics generated through BERTopic

Topic 5 *“Time Management”* residents who improved their time management abilities had a higher pass rate, implying that good time management is a major predictor of success. Topic 6 highlights the importance of residents being knowledgeable about essential procedures such as immunizations, vaccinations, PAP tests, and maintaining vaccination schedules. Mastery of these procedures is crucial for providing comprehensive patient care.

Furthermore, Topic 7, includes *“Teamwork”*, and is strongly correlated with favorable outcomes. Residents who received comments like “team player with excellent communication skills,” “excellent team player!” and “always a team player” passed their certification exams and were designated “PASS”. This shows that teamwork is a crucial indicator of positive performance outcomes.

BERTopic also helps visualize connections between the topics using hierarchical clustering as seen in *Figure 6*, which will help us see the intricately connected topic. For example, note-taking and time management are interconnected, and communication and reasoning in clinical diagnosis are connected; in fact, it also tells that all four of them are connected. Through comprehending these interconnections, the training program may formulate focused treatments to tackle the fundamental concerns comprehensively.

**Figure 6.**
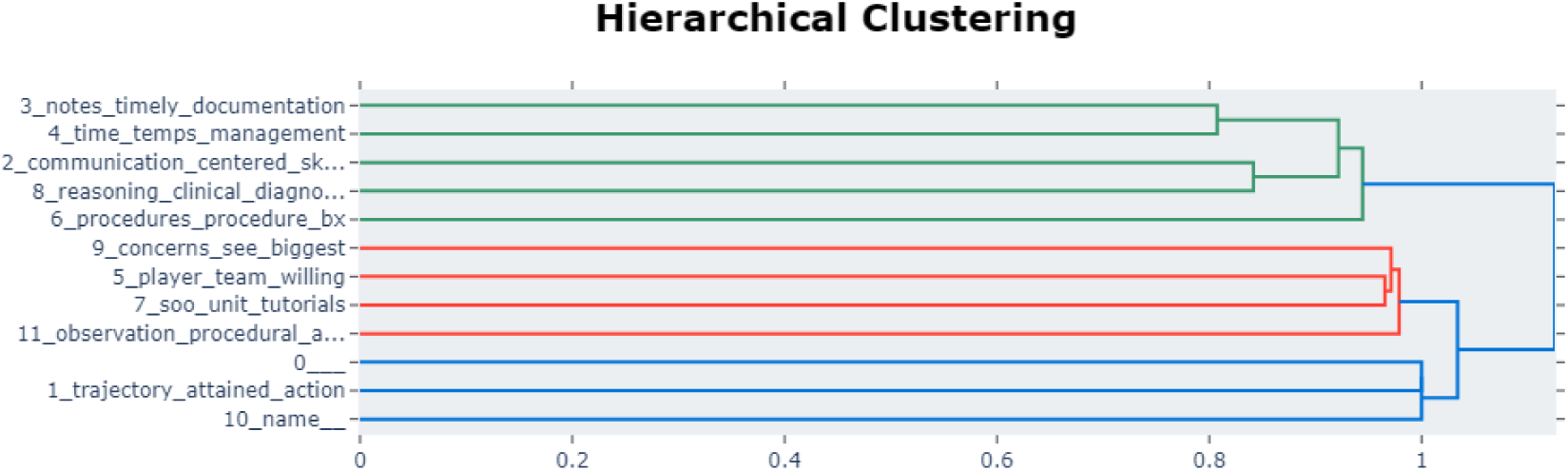
Hierarchical Clustering generated by BERTopic.

## Discussion

The SVM (TF-IDF) model from experiment 3, which utilized a multimodal approach combining qualitative and quantitative data, emerged as the model with the highest performance across experiments 1, 2, and 3, establishing itself as the champion model. This multimodal approach enhanced the prediction performance, comprehensive information and Cross-Domain learning of the model, leveraging the richness of information. Furthermore, the implementation of XAI including SHAP and BERTopic offered insightful analysis of the decision-making process of the model, hence stressing the explainability and transparency of the AI system.

The study’s findings correspond with earlier studies using AI to forecast resident performance. An ML based model using previous data to predict resident performance reached 72% accuracy, showcasing the potential of ML in competency-based education^18^. The critical role of narrative feedback in decision-making is demonstrated by the ability of NLP and ML models to predict underperforming residents with 87% accuracy using narrative feedback from workplace-based assessments (WBAs)^8^. Additionally, classification and regression tree methods identified specific keywords in evaluation reports signaling residents at risk of failure, with precision rates of 23.3 and 23.4%^17^. The enhanced accuracy of our multi-modal technique implies that combining diverse data sources provides a more complete analysis than past studies depending on just one data type. Moreover, our use of XAI provides a better understanding of model decision-making, thereby differentiating our work.

Important business insights can be obtained by directly relating the logical semantic linkages found by BERTopic to the status of the residents. For instance, recognizing that team player traits are associated with successful certification results enables organizations to stress these traits in the procedures of selection, training and assessment. We provide a thorough knowledge of the elements influencing model predictions by using BERTopic to contextualize these insights within natural language explanations and SHAP to determine the significance of input variables.

SHAP and BERTopic integration improves the explainability of our machine learning model and aligns the predictions with business concepts, therefore confirming the practical applicability of our results. The results of the model are guaranteed to be not only technically sound but also relevant and useful for stakeholders with this dual approach.

One Potential reason for the inability of the AI model to flag a few of the cases of the residents that needed intervention could be because their features appeared favourable, which led the AI model to predict otherwise. This inability to flag the struggling residents highlights the challenges of AI (data-driven models), which solely depend on data. These AI model’s misjudgments exemplify the difficulty of training models with only data, as they may need to learn the nuanced contextual elements that physicians identify.

In the subsequent phase a thorough testing is necessary to ensure the AI model’s reliability. As part of this process, we plan to implement testing on a new batch of 41 residents from Fall 2023 data. This will let us evaluate how well the SVM model from experiment 3 handles the new batch. An expert will review the model’s predictions. This testing is a key step before real-world use. The insights we gain will guide future improvements in the next phase.

An AI model might be a useful first screening tool for the training program; after that, the expert clinician could use their knowledge to fine-tune the evaluation process and administer the appropriate intervention. This collaborative approach to flagging can strengthen the early identification of at-risk residents. Consequently, individuals will receive assistance at the precise moment they require it.

Despite promising results, these techniques have limits. First, a relatively small and skewed sample of PGY 1 residents that might compromise generalizability of the model. Our study had a limited dataset of 1,382 data points. Although we attempted to mitigate potential bias by synthetically generating additional data using SMOTE, relying on authentic data would be more ideal for model accuracy. A second limitation could be restricted vocabulary coverage of the AI models, highlighting the necessity for ongoing updates to the model’s training data to include medical terminology and abbreviations.

Our future research aims to enhance the department’s practical application by translating domain-specific knowledge into a usable tool through techniques like domain-specific language modeling and fine-tune models pre-trained on medical corpora such as Bio-BERT^40^ and Bio-GPT^41^. Additionally, we plan to implement data augmentation techniques like GraphRAG^42^ to further improve the model’s performance.

## Conclusion

The successful application of these technologies holds promise for improving the effectiveness of medical residency interventions and contributes to the broader discourse on integrating AI in medical education and practice. By providing a transparent and data-driven approach to identifying at-risk residents, this research represents a significant step forward in leveraging advanced technologies to support the development of medical professionals. The Multi-modal SVM (TF-IDF) model showed the best performance among all the three experiments and was useful in predicting at-risk residents in the medical residency training program. This research emphasizes the advantages of amalgamating qualitative and quantitative data as well as using Explainable AI methods to give insightful analysis for early identification and intervention of residents in difficulty. This accomplishment highlights how innovative technology can improve the medical training program’s ability to identify at-risk residents. Improving the efficacy of medical residency interventions is just one potential outcome of these technologies’ successful implementation; they also add to the larger conversation about using AI in healthcare. Therefore, additional experimentation with more recent datasets is required to establish it conclusively. Visualization from the Explainable AI, of the model gives educators valuable insights into how the prediction models function. This would increase the transparency and enable them to identify at-risk students earlier and implement effective interventions that support their success. This research marks a significant advancement in modern technology to aid in training medical professionals by offering a data-driven method of identifying at-risk residents.

## Data Availability

The data utilized in this study are not publicly available and will not be shared.

## Disclosure of interest

The authors report there are no relevant financial or non-financial competing interests to report.

## Biographical note

1. Abhisht Makarand Joshi holds a Master degree in Computer Science - Applied AI from the University of Ottawa.
2. Pouria Mortezaagha, holds a Master’s degree in System science and engineering from the University of Ottawa.
3. Dr. Diana Inkpen, P.Eng. holds a PhD., From the University of Toronto.
4. Dr Edward Seale, holds MDCM, CCFP(EM), FCFP.
5. Dr. Douglas Archibald, holds a Ph.D from the University of Ottawa.
6. Dr. Kendall Noel, holds MDCM, FCFP, MEd (Organizational Studies).
7. Dr. Arya Rahgozar holds a PhD. From the University of Ottawa.

## Footnotes

a Generative Pre-trained Transformer

b CFPC: College of Family Physicians of Canada

c We used SMOTE^31^ to deal with the imbalance, please refer to methods section for more details.

d https://github.com/abhishtjoshi/Medical-Residency-Lib-Parameters

e Tri-Council Policy Statement

## Notes

### Competing Interest Statement

The authors have declared no competing interest.

### Funding Statement

This study did not receive any funding.

### Author Declarations

The project's scope was reviewed by the Office of Research Ethics and Integrity at the University of Ottawa and was determined to fall within Article 2.5 of the TCPS 2 and was therefore deemed exempt from further review.

### Summary of Updates

Refined the manuscript, specially focused on improving the discussion section.

